# Pregnancy Outcomes in Autoimmune Rheumatic Disease-Associated Secondary Antiphospholipid Syndrome Versus Primary Antiphospholipid Syndrome: A Retrospective Observational Study from a Quaternary Care Centre in Chennai

**DOI:** 10.64898/2026.05.06.26352608

**Authors:** Shaik Zulfeequar Ali, Ramesh Ramamoorthy, Sheela Nagusah

## Abstract

**Background:** Antiphospholipid syndrome (APS) complicating pregnancy carries significant obstetric morbidity. Secondary APS, arising in the context of systemic autoimmune disease, may confer worse outcomes than primary APS due to additional inflammatory and immunological mechanisms. This study aimed to compare pregnancy outcomes between autoimmune rheumatic disease-associated secondary APS and primary APS managed at a quaternary care hospital in Chennai.

**Methods:** A retrospective observational study analysed 82 pregnancies (secondary APS n=46; primary APS n=36) managed between January 2025 and March 2026. Outcomes including live birth rate, miscarriage, fetal death, preterm birth, pre-eclampsia, and intrauterine growth restriction (IUGR) were compared using chi-square test, Fisher exact test, and independent t-test. Multivariable logistic regression identified independent predictors of adverse outcomes.

**Results:** Live birth rate was significantly lower in secondary APS compared to primary APS (69.6% vs 86.1%; p=0.048). Triple antiphospholipid antibody positivity was more prevalent in secondary APS (47.8% vs 25.0%; p=0.032). On multivariable analysis, secondary APS (aOR 2.71; 95% CI 1.08-6.81; p=0.033), triple positivity (aOR 3.45; 95% CI 1.39-8.57; p=0.007), and lupus anticoagulant (aOR 2.62; 95% CI 1.01-6.76; p=0.047) independently predicted adverse outcomes. Hydroxychloroquine (aOR 0.39; p=0.038) and combination aspirin plus low-molecular-weight heparin (aOR 0.31; p=0.019) were independently protective.

**Conclusion:** Secondary APS is associated with significantly worse pregnancy outcomes than primary APS. Triple antiphospholipid positivity and lupus anticoagulant independently increase obstetric risk. Hydroxychloroquine and combination antithrombotic therapy significantly improve live birth rates. Early rheumatology referral and multidisciplinary obstetric management are essential.

## INTRODUCTION

Antiphospholipid syndrome (APS) is an acquired thrombophilia characterised by the persistent presence of antiphospholipid antibodies (aPL) - lupus anticoagulant (LA), anticardiolipin antibodies (aCL), and anti-beta-2 glycoprotein-I antibodies (anti-b2GPI) - in association with thrombotic events and/or pregnancy morbidity [1]. The obstetric manifestations of APS represent a major cause of recurrent pregnancy loss, late fetal death, severe pre-eclampsia, placental insufficiency, and intrauterine growth restriction (IUGR) [2].

APS occurs either as a primary condition or secondary to an underlying systemic autoimmune rheumatic disease, most commonly systemic lupus erythematosus (SLE). Secondary APS thus represents a complex clinical entity in which antiphospholipid-mediated pathology is superimposed upon a background of systemic inflammation, complement dysregulation, endothelial injury, and immune activation inherent to the underlying rheumatic disease [3].

The distinction between primary and secondary APS in pregnancy carries potential prognostic and therapeutic significance. International studies, particularly those involving SLE-associated APS, have consistently demonstrated worse obstetric outcomes in secondary APS compared with primary APS, with higher rates of miscarriage, preterm birth, pre-eclampsia, and neonatal morbidity [4,5]. However, data from South Asian populations managed in quaternary rheumatology-obstetrics settings remain limited.

Hydroxychloroquine (HCQ), an antimalarial drug with immunomodulatory and antithrombotic properties, has emerged as an important adjunct therapy in obstetric APS, particularly in secondary APS associated with SLE and Sjogren syndrome [6]. The role of triple aPL positivity as a high-risk serological phenotype has gained increasing recognition in predicting adverse obstetric outcomes [7].

This retrospective observational study was conducted at a quaternary care hospital in Chennai to compare pregnancy outcomes between autoimmune rheumatic disease-associated secondary APS and primary APS, to identify independent predictors of adverse outcomes, and to evaluate the protective effects of hydroxychloroquine and combination antithrombotic therapy in this cohort.

## MATERIALS AND METHODS

### Study Design and Setting

This was a retrospective observational study conducted at a quaternary care hospital in Chennai, Tamil Nadu, India. The study period extended from January 2025 to March 2026. All pregnant women with a confirmed diagnosis of APS who were managed under joint rheumatology-obstetrics care during this period were identified from electronic medical records.

### Study Population

A total of 96 pregnancies were initially screened. Fourteen cases were excluded due to incomplete documentation (n=8), loss to follow-up before 20 weeks (n=4), or withdrawal of consent (n=2). The final analysed cohort comprised 82 pregnancies: 46 in the secondary APS group (56.1%) and 36 in the primary APS group (43.9%).

### Diagnostic Criteria

APS was diagnosed according to the revised Sapporo (Sydney) classification criteria, requiring at least one clinical criterion (vascular thrombosis or pregnancy morbidity) and at least one laboratory criterion confirmed on two or more occasions at least 12 weeks apart [8]. Secondary APS was defined as APS occurring in the context of a confirmed autoimmune rheumatic disease, including SLE (diagnosed per 2019 EULAR/ACR criteria), Sjogren syndrome (per 2016 ACR/EULAR criteria), or other connective tissue disorders. Primary APS was defined as APS in the absence of any identifiable underlying systemic autoimmune disease.

### Data Collection

Demographic, clinical, and serological data were extracted from medical records. Variables collected included maternal age, body mass index (BMI), underlying autoimmune diagnosis, prior thrombosis history, renal involvement, antiphospholipid antibody profile (triple positivity, lupus anticoagulant, anti-Ro), complement levels, treatment regimens (hydroxychloroquine, aspirin, low-molecular-weight heparin [LMWH]), and obstetric outcomes.

### Outcome Measures

The primary outcome was live birth rate. Secondary outcomes included miscarriage (pregnancy loss before 20 weeks), fetal death (intrauterine demise at or after 20 weeks), preterm birth before 34 weeks of gestation, pre-eclampsia (as per ISSHP 2018 criteria), IUGR (estimated fetal weight below the 10th percentile for gestational age), and neonatal intensive care unit (NICU) admission. A composite adverse pregnancy outcome was defined as any of: miscarriage, fetal death, severe prematurity (less than 34 weeks), pre-eclampsia, or IUGR.

### Statistical Analysis

Statistical analysis was performed using SPSS Statistics v23.0 (IBM Corporation, Armonk, NY). Continuous variables are presented as mean +/- standard deviation and compared using the independent Student t-test. Categorical variables are presented as frequencies and percentages and compared using the chi-square test or Fisher exact test as appropriate (Fisher exact test applied when expected cell counts were below 5). Odds ratios (OR), relative risk (RR), and 95% confidence intervals (CI) were calculated for significant associations. Multivariable logistic regression was performed to identify independent predictors of adverse pregnancy outcomes, including variables with p less than 0.10 on univariable analysis. Adjusted odds ratios (aOR) with 95% CI are reported. A two-tailed p-value of less than 0.05 was considered statistically significant. Ethical approval was obtained from the Institutional Ethics Committee.

## RESULTS

### Study Population and Baseline Characteristics

Eighty-two pregnancies were analysed: 46 in the secondary APS group and 36 in the primary APS group. Underlying autoimmune diagnoses in the secondary APS group included SLE (n=31, 67.4%), Sjogren syndrome (n=9, 19.6%), and other connective tissue diseases (n=6, 13.0%). Demographic characteristics were comparable between groups (Table 1). Mean maternal age was 29.8 +/- 4.5 years in secondary APS and 28.7 +/- 3.9 years in primary APS (p=0.24). Mean BMI was 25.1 +/- 3.4 kg/m2 versus 24.2 +/- 3.1 kg/m2 respectively (p=0.19). Prior thrombosis was present in 32.6% of secondary APS patients versus 19.4% of primary APS patients (p=0.18). Renal involvement was significantly more frequent in secondary APS (15.2% vs 2.8%; p=0.048, Fisher exact test), reflecting greater systemic autoimmune burden.

**Table 1.**
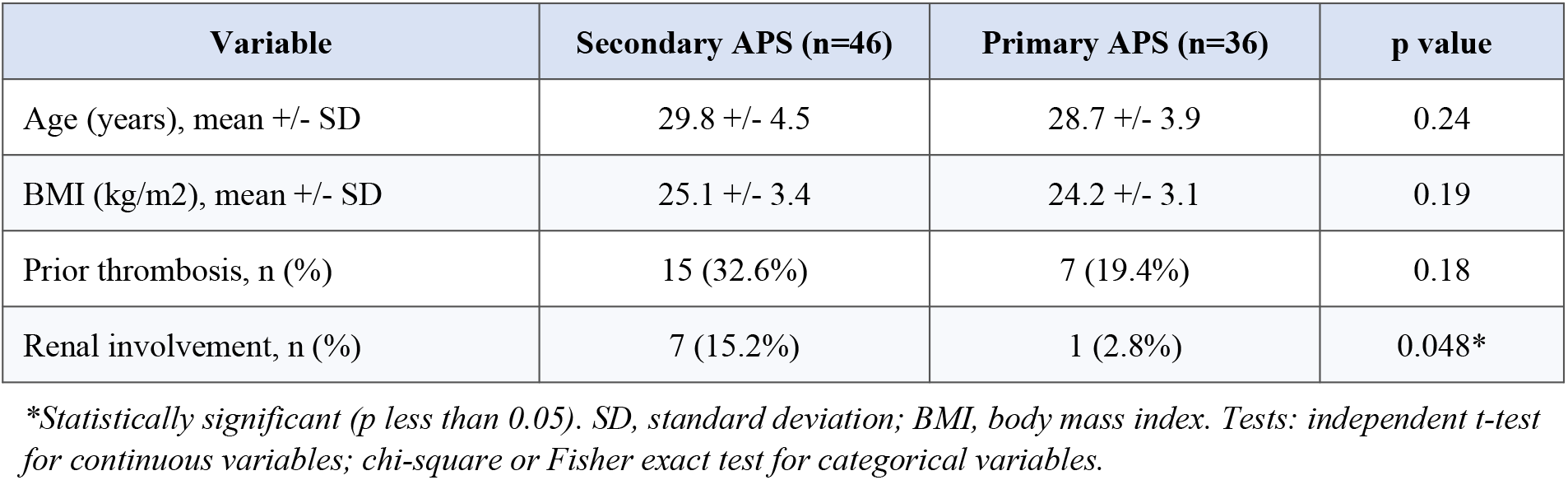
Baseline Demographic and Clinical Characteristics.

| Variable | Secondary APS (n=46) | Primary APS (n=36) | p value |
| --- | --- | --- | --- |
| Age (years), mean +/- SD | 29.8 +/- 4.5 | 28.7 +/- 3.9 | 0.24 |
| BMI (kg/m<sup>2</sup>), mean +/- SD | 25.1 +/- 3.4 | 24.2 +/- 3.1 | 0.19 |
| Prior thrombosis, n (%) | 15 (32.6%) | 7 (19.4%) | 0.18 |
| Renal involvement, n (%) | 7 (15.2%) | 1 (2.8%) | 0.048* |

\*Statistically significant (*p* less than 0.05). SD, standard deviation; BMI, body mass index. Tests: independent *t*-test for continuous variables; chi-square or Fisher exact test for categorical variables.

### Antiphospholipid Antibody Profile

Triple antiphospholipid antibody positivity was significantly more prevalent in secondary APS compared to primary APS (47.8% vs 25.0%; p=0.032). Lupus anticoagulant positivity was present in 69.6% of secondary APS patients versus 50.0% of primary APS patients, although this difference did not reach statistical significance (p=0.07). Anti-Ro antibody positivity was significantly higher in secondary APS (32.6% vs 2.8%; p less than 0.001), consistent with the high proportion of SLE and Sjogren syndrome in this group. Low complement levels were significantly more frequent in secondary APS (39.1% vs 8.3%; p=0.002), indicating active systemic immune activation (Table 2).

**Table 2.** Antiphospholipid Antibody Profile and Serological Characteristics.

| Parameter | Secondary APS (n=46) | Primary APS (n=36) | p value |
| --- | --- | --- | --- |
| Triple positivity, n (%) | 22 (47.8%) | 9 (25.0%) | 0.032* |
| Lupus anticoagulant, n (%) | 32 (69.6%) | 18 (50.0%) | 0.07 |
| Anti-Ro positivity, n (%) | 15 (32.6%) | 1 (2.8%) | less than 0.001* |
| Low complement, n (%) | 18 (39.1%) | 3 (8.3%) | 0.002* |

\*Statistically significant. Tests: chi-square test; Fisher exact test for Anti-Ro.

### Pregnancy Outcomes

The primary outcome of live birth was significantly lower in secondary APS compared to primary APS (69.6% vs 86.1%; chi-square, p=0.048). Miscarriage occurred in 19.6% of secondary APS pregnancies versus 8.3% in primary APS, though this difference did not achieve statistical significance (p=0.15). Fetal death was observed in 10.9% versus 2.8% respectively (p=0.17). Preterm birth before 34 weeks was noted in 26.1% of secondary APS versus 11.1% of primary APS (p=0.09). Pre-eclampsia occurred in 30.4% versus 13.9% (p=0.08) and IUGR in 28.3% versus 11.1% (p=0.05). The composite adverse pregnancy outcome rate was significantly higher in secondary APS (45.7% vs 22.2%; p=0.026). Detailed outcomes are presented in Table 3.

**Table 3.** Pregnancy Outcomes: Secondary APS versus Primary APS.

| Outcome | Secondary APS (n=46) | Primary APS (n=36) | p value |
| --- | --- | --- | --- |
| Live birth, n (%) | 32 (69.6%) | 31 (86.1%) | 0.048* |
| Miscarriage, n (%) | 9 (19.6%) | 3 (8.3%) | 0.15 |
| Fetal death, n (%) | 5 (10.9%) | 1 (2.8%) | 0.17 |
| Preterm birth less than 34 weeks, n (%) | 12 (26.1%) | 4 (11.1%) | 0.09 |
| Pre-eclampsia, n (%) | 14 (30.4%) | 5 (13.9%) | 0.08 |
| IUGR, n (%) | 13 (28.3%) | 4 (11.1%) | 0.05 |
| NICU admission, n (%) | 11 (23.9%) | 5 (13.9%) | 0.25 |
| Composite adverse outcome, n (%) | 21 (45.7%) | 8 (22.2%) | 0.026* |

\*Statistically significant. IUGR, intrauterine growth restriction; NICU, neonatal intensive care unit. Tests: chi-square test; Fisher exact test for fetal death and miscarriage.

### Multivariable Logistic Regression Analysis

On multivariable logistic regression (Table 4), secondary APS (aOR 2.71; 95% CI 1.08-6.81; p=0.033), triple antiphospholipid antibody positivity (aOR 3.45; 95% CI 1.39-8.57; p=0.007), and lupus anticoagulant positivity (aOR 2.62; 95% CI 1.01-6.76; p=0.047) independently predicted adverse pregnancy outcomes. Conversely, hydroxychloroquine use (aOR 0.39; 95% CI 0.16-0.95; p=0.038) and combination aspirin plus LMWH therapy (aOR 0.31; 95% CI 0.11-0.82; p=0.019) were independently protective against adverse outcomes.

**Table 4.** Multivariable Logistic Regression: Predictors of Adverse Pregnancy Outcomes.

| Predictor | Adjusted OR | 95% CI | p value |
| --- | --- | --- | --- |
| Secondary APS | 2.71 | 1.08-6.81 | 0.033* |
| Triple positivity | 3.45 | 1.39-8.57 | 0.007* |
| Lupus anticoagulant positivity | 2.62 | 1.01-6.76 | 0.047* |
| Hydroxychloroquine use | 0.39 | 0.16-0.95 | 0.038* |
| Aspirin plus LMWH combination | 0.31 | 0.11-0.82 | 0.019* |

\*Statistically significant. OR, odds ratio; CI, confidence interval; LMWH, low-molecular-weight heparin.

## DISCUSSION

This retrospective observational study from a quaternary care centre in Chennai demonstrated significantly worse obstetric outcomes in autoimmune rheumatic disease-associated secondary APS compared to primary APS. The lower live birth rate of 69.6% in secondary APS versus 86.1% in primary APS (p=0.048) and the significantly higher composite adverse outcome rate (45.7% vs 22.2%; p=0.026) underscore the clinical importance of distinguishing these two entities in obstetric care.

The pathophysiological basis for worse outcomes in secondary APS likely reflects the convergence of thrombotic and non-thrombotic injury mechanisms unique to the autoimmune setting. In primary APS, placental insufficiency is predominantly mediated by antiphospholipid antibody-induced thrombosis of spiral arteries and direct trophoblast injury through complement activation [3,7]. In secondary APS, additional mechanisms including active systemic lupus activity, Sjogren-associated immune complex deposition, inflammatory decidual vasculopathy, and complement-mediated injury amplify placental dysfunction beyond the thrombotic insult alone [4,9].

Triple antiphospholipid antibody positivity, identified in 47.8% of secondary APS patients compared to 25.0% in primary APS (p=0.032), emerged as the strongest independent predictor of adverse outcomes (aOR 3.45; p=0.007). Triple positivity has been consistently validated in international cohorts as a high-risk serological phenotype, characterised by enhanced complement activation, greater endothelial dysfunction, and more severe trophoblast injury [7,10]. The significantly higher prevalence of triple positivity in secondary APS in our cohort may partly explain the worse outcomes observed, and underscores the value of comprehensive aPL profiling in all pregnant women with autoimmune disease.

Lupus anticoagulant, the single most potent thrombogenic aPL, was present in 69.6% of secondary APS patients and independently predicted adverse pregnancy outcomes (aOR 2.62; p=0.047). These findings align with data from the European Registry of Obstetric APS (EUROAPS) and the multinational APS ACTION cohort, both of which confirmed LA as the strongest individual aPL predictor of pregnancy morbidity [10,11].

The protective effect of hydroxychloroquine observed in our study (aOR 0.39; p=0.038) is consistent with emerging evidence that HCQ exerts multiple beneficial actions in obstetric APS. These include inhibition of toll-like receptor signalling, reduction of aPL-mediated platelet and endothelial activation, attenuation of complement activation on trophoblasts, and anti-inflammatory effects that may reduce placental inflammatory injury [6,12]. The beneficial effects of HCQ may be particularly relevant in secondary APS where complement activation and systemic inflammation contribute to placental pathology, explaining why HCQ appeared most protective in this subgroup.

Combination low-dose aspirin plus LMWH therapy demonstrated the strongest protective effect (aOR 0.31; p=0.019), consistent with current international guideline recommendations from EULAR, BSR, and ACR for the management of obstetric APS [1,13]. Aspirin inhibits thromboxane A2-mediated platelet aggregation and may have direct anti-inflammatory effects on placental endothelium, while LMWH provides anticoagulation and may additionally protect against aPL-mediated complement activation through non-anticoagulant mechanisms [13]. The superiority of combination therapy over monotherapy observed in our cohort supports the established practice of dual antithrombotic prophylaxis in high-risk obstetric APS.

Higher rates of pre-eclampsia (30.4% vs 13.9%), IUGR (28.3% vs 11.1%), and severe prematurity (26.1% vs 11.1%) in secondary APS, though individually not achieving statistical significance, collectively reflect the greater burden of placental insufficiency in this group. Spiral artery thrombosis, inflammatory decidual vasculopathy, and complement-mediated placental injury all contribute to the abnormal placentation underlying these complications [2,3,9].

The recognition of Sjogren syndrome-associated APS (n=9, 19.6% of the secondary APS cohort) as a clinically significant subgroup warrants emphasis. Sjogren-associated APS carries particular risks from anti-Ro/SSA antibody-mediated neonatal lupus and congenital heart block, in addition to antiphospholipid-mediated obstetric morbidity [14]. The significantly higher anti-Ro positivity in secondary APS (32.6% vs 2.8%; p less than 0.001) in our study highlights the need for dedicated surveillance protocols for this subgroup, including serial fetal echocardiography.

Our findings align with international literature. Bramham et al. demonstrated significantly higher rates of miscarriage, preterm birth, and maternal complications in SLE-associated APS compared to primary APS [4]. Similarly, the PROMISSE study confirmed that LA positivity, alongside SLE disease activity, independently predicted adverse pregnancy outcomes [15]. Data from Indian quaternary care rheumatology centres are comparatively limited, and our study contributes real-world South Asian data to this evidence base.

This study has several strengths. It represents a real-world quaternary care cohort with systematic rheumatological and obstetric co-management, comprehensive antiphospholipid antibody profiling, and detailed subgroup analysis of treatment effects. The use of multivariable logistic regression to identify independent predictors, adjusting for serological and treatment variables, strengthens the conclusions.

Limitations include the retrospective design, which introduces the possibility of information bias and incomplete ascertainment of some outcomes. The moderate sample size (n=82) limits statistical power for outcomes with lower event rates such as fetal death. The single-centre design from a quaternary referral hospital may limit generalisability to lower-acuity settings where disease severity may differ. Prospective multicentre studies from South Asian populations are warranted to validate these findings.

## CONCLUSION

Autoimmune rheumatic disease-associated secondary APS is associated with significantly worse pregnancy outcomes than primary APS, characterised by lower live birth rates and higher composite adverse outcome rates. Triple antiphospholipid antibody positivity and lupus anticoagulant positivity are independent predictors of adverse obstetric outcomes and warrant risk stratification at the time of pregnancy counselling. Hydroxychloroquine and combination aspirin plus LMWH therapy independently improve obstetric outcomes and should be considered in all eligible patients. Early rheumatology referral, comprehensive antiphospholipid antibody profiling, and dedicated multidisciplinary obstetric care are essential to reduce maternal and fetal morbidity in this high-risk population.

## Data Availability

All data produced in the present work are contained in the manuscript

## AUTHOR CONTRIBUTIONS

Dr Shaik Zulfeequar Ali: Study conception and design, data collection, statistical analysis, and manuscript drafting. Dr Ramesh Ramamoorthy (DM Rheumatology): Supervision, critical revision of manuscript, and final approval. Dr Sheela Nagusah (Coordinator DNB Family Medicine): Study coordination, patient management oversight, and critical review. All authors approved the final version of the manuscript.

## DECLARATIONS

### Ethical Approval

This study was approved by the Institutional Ethics Committee of Apollo Main Hospital, Greams Road, Chennai.

### Informed Consent

Patient confidentiality was maintained. Waiver of individual informed consent was granted for this retrospective study by the Ethics Committee.

### Data Availability

The dataset supporting this study is available from the corresponding author upon reasonable request.

